# Understanding the immunological landscape of England during SARS-CoV2 Omicron variant wave

**DOI:** 10.1101/2022.02.21.22271270

**Authors:** Joseph Shingleton, Steven Dyke, Archie Herrick, Thomas Finnie

## Abstract

Understanding the scale of the threat posed by SARS-CoV2 B.1.1.529, or *Omicron*, variant formed a key problem in public health in the early part of 2022. Early evidence indicated that the variant was more transmissible and less severe than previous variants. As the virus was expected to spread quickly through the population of England, it was important that some understanding of the immunological landscape of the country was developed. This paper attempts to estimate the number of people with good immunity to the Omicron variant, defined as either recent infection with two doses of vaccine, or two doses of vaccine with a recent booster dose. To achieve this, we use a process of iterative proportional fitting to estimate the cell values of a contingency table, using national immunisation records and real-time model infection estimates as marginal values. Our results indicate that, despite the increased risk of immune evasion with the Omicron variant, a high proportion of England’s population had good immunity to the virus, particularly in older age groups. However, low rates of immunity in younger populations may allow endemic infection to persist for some time.

## Introduction

A crucial factor in understanding the public health challenges posed by the spread of SARS-CoV2 B.1.1.529, or *Omicron*, variant is identifying the size of the population likely to be susceptible to the variant. Previous studies have identified that while individuals with two doses of vaccine may be protected against severe disease, the best protection against both infection and severe disease comes from recent booster doses of vaccine [1]. Further, it likely that the Omicron variant may evade immunity acquired from previous infection, particularly from infection with B.1.1.7 (Alpha) and B.1.617.2 (Delta) variants [2, 3].

Vaccination rates in over 12s in the UK are generally high, with 83% having received two doses of either Astra-Zeneca, Moderna or Pfizer vaccines [4]. A rapid roll out of booster vaccines, primarily either Pfizer or Moderna, has ensured that 63% of over 12s have received three doses of vaccine as of 14^th^ of January 2022 [4]. Further, there are high levels of infection acquired immunity in England, following three waves of COVID-19 infections in 2020 and 2021.

This report aims to identify the level of immunity in England across six age groups. To achieve this we consider eight groups of individuals – those who are unvaccinated and have no prior infection, those whose are unvaccinated but have had a prior infection, and those who have received one, two or three vaccines, both with and without prior infection. Further, we separate those with prior infection into ‘recent’ and ‘non-recent’ infections, helping to account for waning immunity. We consider individuals with either three doses of vaccine, or two doses plus recent prior infection, as having good immunity to the Omicron variant.

Finally, we compare the current immunological profile of England with that prior to the wave of Delta variant infections in May 2021. Due to immunological differences between Omicron and Delta variants, we consider good protection to Delta variant to be provided by two doses of vaccine with any prior infection status, or one dose with recent prior infection.

## Methods

The total population with one, two or three doses of vaccine are taken from the National Immunisation Management System (NIMS) dataset [4]. This dataset does not make any distinction between third doses (i.e. full vaccine doses given to clinically vulnerable individuals) and booster doses. The analysis presented in this report does not attempt to account for time since vaccination. We assume all third doses have occurred recently, and offer a good level of protection to Omicron variant. Any other vaccination status without recent prior infection is considered to offer poor protection to Omicron variant.

We estimate the number of people infected with COVID-19 using the UKHSA-Cambridge MRC Real Time Model – a transmission model used to estimate new COVID-19 infections based on a number of different data sources [5]. Infections are further separated into ‘recent’ and ‘non-recent’, defining ‘recent’ infections as those occurring within the last 3 months.

The number of people with no vaccination and the number of people with no prior infection are calculated using Office for National Statistics 2020 mid-year population estimates [6]. In some cases, particularly in the oldest age group, this leads to vaccination rates exceeding one-hundred percent. In such cases, we adjust the total population to give a one-hundred percent vaccination rate.

The total vaccinated and previously infected populations in each age group are used as marginal totals in a contingency table. We use Iterative Proportional Fitting (IPF) to estimate the cell values in each contingency table, giving us the number of people in each vaccination group with recent, non-recent and no prior infection [7].

We calculate 95% confidence intervals for each estimated value assuming a normal approximation with mean equal to the estimate and standard deviation as calculated in [7] using an asymptotic approximation of the covariance matrix. This method permits negative values for the lower confidence interval. Where this occurs we assume the physical value of the lower interval to be zero.

## Results

Figure 1 shows the immunological profile for England across each age group, both as total population (fig 1a) and proportion of population (fig 1b). The analysis indicates that there is a good level of immunity in older age groups, with over 90% of over 65s having either three doses of vaccine or two doses with recent infection. Immunity is also high in the 45-65 age group, with over 80% having good levels of immunity.

**Figure 1:**
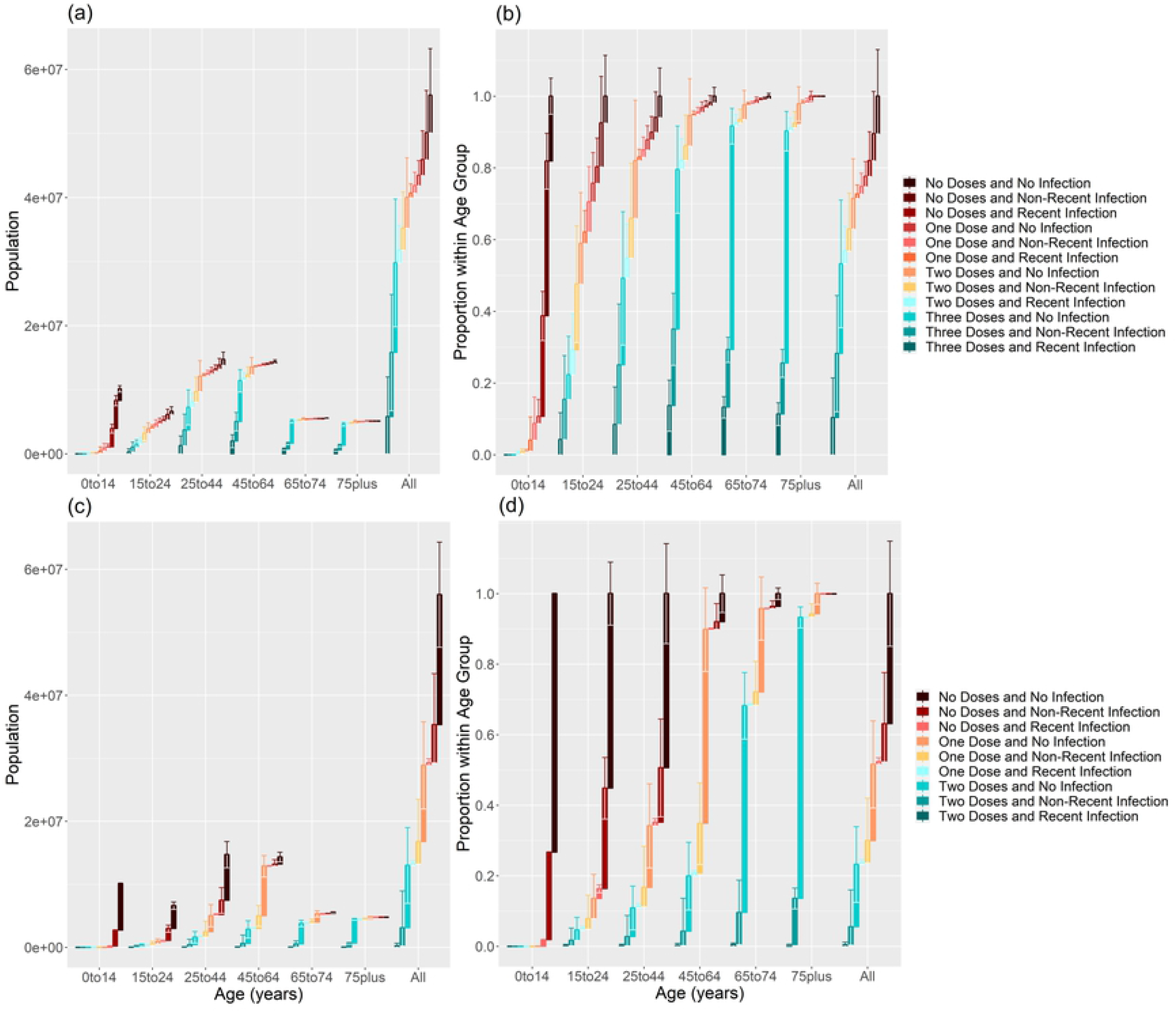
Immunological profiles across six age groups in England. (a) and (b) show the absolute and proportional vaccination and infection profiles as of 12th of January 2022, respectively, while (c) and (d) show the same values using data from 1st of May 2021, prior to the Delta-variant wave of infections. The whiskers at the top of each bar indicate the 95% confidence interval of the estimate.

Immunity levels fall quickly in younger age groups, with only around 50% of 25-45 year olds having had either three doses of vaccine or two doses with recent prior infection, falling to around 30% in 15-25 year olds. Immunity rates in younger children are very low.

Figure 1b and 1c show the same analysis performed on data prior to the wave of Delta variant infections starting in May 2021. Note that the colours have been changed in this plot to highlight the increased level of protection provided by two doses of vaccine, compared with the Omicron variant. Immunity in over 75s is currently comparable to the pre-Delta period. Currently, there are significantly higher levels of immunity in younger age groups compared to the pre-Delta period.

## Discussion

The current immunological landscape in England is varied. There is a good level of protection against infection and severe disease in older populations – which will help to reduce the pressures on healthcare systems. However, the lower rates of immunity in working age people is an important concern. Widespread infection in these groups, regardless of severity, will put considerable stress on businesses and services as a result of the legal requirement to self isolate upon returning a positive COVID-19 test. Further, the low rates of immunity in younger children will likely lead to uncontrolled spread of Omicron variant in schools, which may go on to seed community outbreaks.

The analysis presented in this report goes some way to highlighting the groups most at risk of widespread infection, and raises concerns about low immunity rates in younger populations. However, there are some limitations to the analysis which should be considered.

The IPF analysis is a purely statistical method, with no mechanistic aspect. As such, the analysis does not account for the protection against disease afforded by vaccination. This means that prior infections are likely under-estimated in unvaccinated groups. This effect is mitigated somewhat by the analysis focussing mainly on broad groups of ‘good’ and ‘poor’ immunity, as unvaccinated individuals are considered as having poor immunity, regardless of prior infection status.

The analysis does not consider time since vaccination as a factor influencing immunity. Once again this is mitigated by the broad characterisation of ‘good’ and ‘poor’ immunity, and the assumption that all third doses are immunologically recent. While a recent second dose of vaccine likely confers more immunity than a non-recent one, we consider both to provide ‘poor’ immunity to Omicron variant. Similarly, we assume that two doses of mRNA vaccine (Moderna or Pfizer) plus a booster dose offers a similar level of protection as two doses of Astra-Zeneca vaccine plus a booster dose of mRNA vaccine.

The interpretation of this analysis is based on a number of assumptions on the level of immunity offered by vaccination status and prior infection. Further work is necessary to properly quantify this immunity, and more accurately estimate the level of protection present in the population. Similarly, it would be useful to consider immunity in two parts – first as protection against infection and second as a protection against severe disease. Both outcomes present unique challenges, particularly in healthcare settings in which both widespread infections in staff, and increased numbers of severe COVID-19 infections, are resulting in considerable system stresses.

## Data Availability

Vaccination data are from the National Immunisation Managment Service (NIMS) database and are not publically avaialable. This is currently managed by NHS England, and appropriate access can be required via https://digital.nhs.uk/coronavirus/vaccinations/training-and-onboarding/point-of-care/national-immunisation-management-service-nims-app. Estimates for COVID-19 infections are taken from the UKHSA-Cambridge COVID-19 infection model. This is currently managed by both the UK Health Security Agency and Cambridge MRC Bio Statistics Unit. The data are not publically available.

